# Use of stacked proportional bar graphs (“Grotta bars”) to visualize functional outcome distributions in observational neurology research

**DOI:** 10.1101/2024.07.08.24310003

**Authors:** Meghan R. Forrest, Tracey L. Weissgerber, Emma S. Lieske, Elena Tamayo Cuartero, Elena Fischer, Lydia Jones, Marco Piccininni, Jessica L. Rohmann

**Author notes:** **Correspondence:** Meghan R. Forrest, MSc, Institute of Public Health, Charité – Universitätsmedizin Berlin, Charitéplatz 1, 10117 Berlin, Germany.

## Abstract

**Background and Objectives:** Stacked proportional bar graphs (nicknamed “Grotta bars”) are commonly used to visualize functional outcome scales in stroke research and are also used in other domains of neurological research. In observational studies that present adjusted effect estimates, Grotta bars can mislead readers if they show unadjusted, confounded comparisons. In a sample of recent observational neurology studies with confounding-adjusted effect estimates, we aimed to determine the frequency with which Grotta bars were used to visualize functional outcomes and how often unadjusted Grotta bars were presented without an accompanying adjusted version. We also assessed the methods used to generate adjusted Grotta bars.

**Methods:** In this meta-research study, we systematically examined all observational studies published in the top 15 Clinical Neurology journals between 2020-2021 with an ordinal functional outcome and confounding-adjusted effect estimate. We determined whether at least one comparison using Grotta bars was present, whether the visualized comparisons were adjusted, and which adjustment strategies were applied to generate these graphs.

**Results:** 250 studies met all inclusion criteria. Of these, 93 (37.2%) used Grotta bars to depict functional outcome scale distributions, with 73 (81.7%) presenting only Grotta bars without model-based adjustment. Amongst the 17 studies that presented Grotta bars adjusted using a model, the adjustment strategies included propensity score matching (n=10; 58.8%), regression (n=6; 35.3%), and inverse probability weighting (n=1; 5.9%). Most studies with Grotta bars (n=87; 87.9%) were stroke studies.

**Discussion:** Grotta bars were most often used in stroke research within our sample. Papers that present adjusted associations for functional outcomes commonly showed only unadjusted Grotta bars, which alone may be misleading for causal questions. In observational research, Grotta bars are most informative if an adjusted version, aligning with adjusted effect estimates, is presented directly alongside the unadjusted version. Based on our findings, we offer recommendations to help authors generate informative Grotta bars and facilitate correct interpretation for readers.

## Introduction

Stacked proportional bar graphs are commonly used to present functional outcomes that quantify global physical functional ability on an ordinal scale.^1^ In the stroke field, these visualizations acquired the nickname “Grotta bars” after James C. Grotta and colleagues used them to compare functional outcome scales between intervention groups in the 1996 rt-PA trial.^2,3^ Thereafter, Grotta bars became standard in the reporting of clinical stroke studies.^1^ These graphs are especially attractive for presenting ordinal outcomes, such as the 7-point modified Rankin Scale (mRS).^1,4^ They allow the visual representation of each level of the scale,^1,3^ which is preferable to discarding information by improperly dichotomizing ordinal outcomes (e.g., “good outcome” versus “poor outcome”).^4–8^ This allows Grotta bars to granularly depict the difference (“shift”) of the full mRS distribution between the exposure groups.^3,4^ These characteristics make Grotta bars particularly useful to researchers, policy and decision-makers, clinicians, and patients when interpreting study results.^5,8,9^

In well-conducted, large randomized controlled trials, any visible shift in the distribution of the outcome between exposure groups has a direct causal interpretation that corresponds to the effect of the intervention. The interpretation of Grotta bars in observational studies is more complex. Observed shifts cannot be interpreted causally unless the authors rigorously applied suitable causal inference methods.^10^ Unadjusted Grotta bars showing only observed data (e.g., without confounding control) may mislead readers if interpreted causally.^4^

A recent study using data from a large registry illustrated this problem by examining the relationship between being “discharged home” versus “discharged elsewhere” and functional outcomes following a stroke or transient ischemic attack.^4^ While the unadjusted results appeared to show a dramatic difference, evidence of any association between the exposure and outcome disappeared once inverse probability weighting was applied to control for confounding. The authors generated two Grotta bar graphs depicting the unadjusted and adjusted outcome comparisons (**Figure 1**). The large shift seen in the unadjusted Grotta bars was greatly attenuated after adjustment for confounding, which aligned with the computed unadjusted and adjusted common odds ratios.^4^

**Figure 1.**
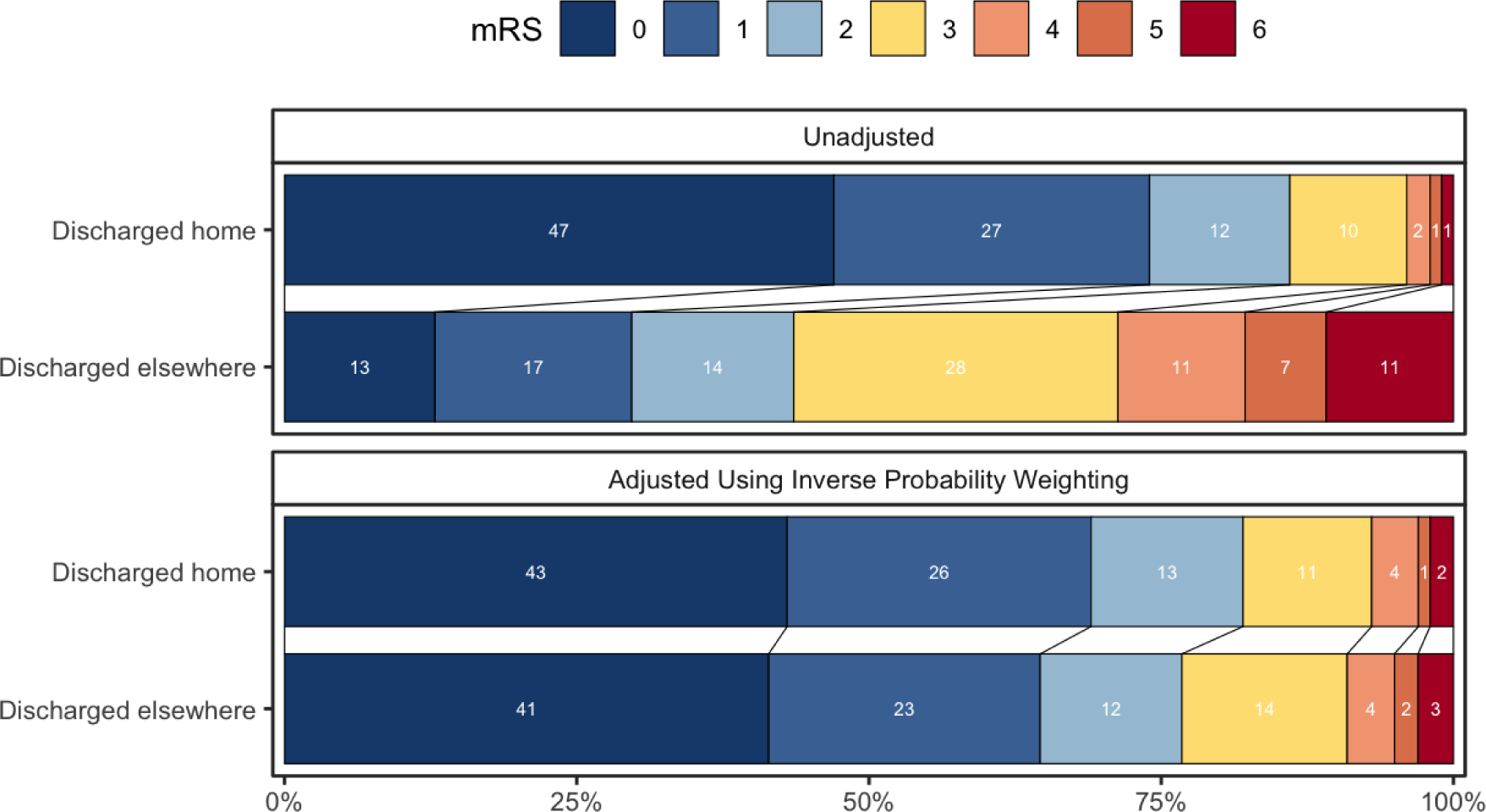
Depiction of example Grotta bars. Grotta bars depicting the distributions of mRS scores for ischemic stroke patients by discharge type (discharged home or discharged elsewhere) without confounding adjustment (top) and with confounding adjustment using inverse probability weighting (bottom). Figure adapted from Rohmann et al. (Figure 2).^4^

Researchers can use several different methods to generate confounding-adjusted effect estimates^10^ and Grotta bars.^4^ In observational neurology studies, confounding control is frequently performed using traditional outcome regression or other methods such as propensity-score-based adjustment (including weighting and matching). A few papers in the stroke literature have presented confounding-adjusted Grotta bars to accompany their confounding-adjusted effect estimates,^11,12^ although this is uncommon.

While we know this potential problem is relevant to clinical literature about stroke populations, it may also be relevant to research examining other neurological diseases that also use ordinal functional outcomes. In observational neurological studies that adjust for confounding, we aim to assess how often Grotta bars are used to visualize ordinal functional outcomes, how often adjusted effect estimates are accompanied by adjusted Grotta bars, and which statistical methods are used to generate confounding-adjusted Grotta bars.

## Methods

We systematically reviewed observational studies in top clinical neurology journals. No ethics approval was required for this study of published literature. All protocols underwent feasibility testing and were pre-registered on our Open Science Framework (RRID:SCR_003238) repository which also contains full protocols, protocol modifications, and a full abstraction table: https://osf.io/w78mh/. Data and code can be accessed on our GitHub repository: https://github.com/meghanrforrest/stackedproportionalbargraph-meta-research-study.

### Journal Screening

We retrieved a list of all journals indexed in the 2021 Science Citation Index-Expanded classification of the Journal Citation Reports (RRID:SCR_017656) Clinical Neurology category and then sorted this list according to the 2021 Journal Impact Factor. Starting with the highest impact factor, two independent reviewers (MRF and ESL) evaluated whether each journal (1) published articles in English, and (2) published full-length original research. There were no discrepancies between reviewers. The top 15 journals meeting these inclusion criteria were incorporated into our search strategy **(Supplement: Table 1)**.

### Search Strategy

Our search strategy was developed in consultation with an information specialist (LJ). All journals included in our search strategy were indexed in PubMed (RRID:SCR_004846) in 2020 and 2021. We retrieved all bibliographic records from PubMed, which were published in the journals identified for inclusion either in print or electronically between 2020-2021, and applied a validated search filter to identify observational research (sensitivity: 92.4%, specificity: 79.7%)^13^. The full search strategy is detailed in **Supplement: Table 2**.

### Inclusion and Exclusion Criteria

We included all articles in our sample which (1) were full-length, original research; (2) were written in English; (3) had human participants; (4) were observational studies that aimed to identify a cause-effect relationship; (5) had a functional outcome as a dependent variable of any analysis within the study; and (6) contained a confounding-adjusted effect estimate for the functional outcome.

We excluded each article from our study that was (1) short-form research, such as a brief report, research letter, a paper without a Background, Methods, Results, or Discussion section, or not original research; (2) published in a language other than English; (3) an animal study; (4) a prediction model development study, systematic review, or non-observational research; (5) lacking a functional outcome as a dependent variable of the analysis; or (6) without an effect estimate for the functional outcome, or contained only effect estimates which were not adjusted for confounding.

### Article Screening

Both title/abstract screening and full-text screening were conducted using Rayyan QCRI (RRID:SCR_017584). All bibliographic records retrieved from the search strategy first underwent title/abstract screening by two independent reviewers (MRF and either ESL or ETC). If reviewers were unsure or disagreed about whether an article met all inclusion criteria, the article proceeded to full-text screening, during which two independent reviewers (MRF and either EF, ESL, or ETC) evaluated its qualification for inclusion. All four reviewers (MRF, EF, ESL, ETC) resolved discrepancies arising from full-text screening via group consensus. If consensus could not be achieved, a fifth reviewer (JLR) was consulted for arbitration.

### Abstraction

Each article that met all inclusion criteria was abstracted by two independent abstractors (MRF and either EF, ESL, or ETC). Abstractors first confirmed that the article met all inclusion criteria.

The following data were abstracted for analysis:

- Characteristics of each paper (study design, population, functional outcome)
- Presence of Grotta bars visualizing the functional outcome (yes/no)
- Presence of figures that were not Grotta bars to visualize the functional outcome (yes/no)
- For each Grotta bars graph: was a model-based adjustment method (e.g., ordinal regression, propensity score matching, inverse probability weighting) applied to create the bars? If yes, which method was applied?
- For each Grotta bars graph: is it stratified?

### Statistical Analysis

In this descriptive analysis, summary statistics were calculated using R (version 4.2.2) and RStudio (version 2023.03.0+386). Frequencies and percentages were used to describe all categorical variables.

Grotta bars that were generated using a model-based adjustment method were categorized differently than those that were stratified. While stratification is a valid strategy to remove confounding, it only removes confounding related to the stratifying variable(s). Grotta bars comparing exposure groups stratified by only a few variables, e.g. only by sex, are likely not sufficient to address the threat of unmeasured confounding. For this reason, we considered stratified Grotta bars as a standalone category.

Per our inclusion criteria, we sought to include studies with causal aims. Adjustment for confounding is a characteristic of causal research^14^; however, ascertaining whether studies have causal intentions is often not straightforward. The language authors use can be unreliable when determining whether a study has causal aims.^15,16^ Furthermore, the statistical methods applied in causal and predictive studies often overlap, with interpretations that are frequently conflated in health research.^17,18^

Our complete “likely causal” sample contained studies that appeared to the review team as having potential causal aims. Some of these articles also had characteristics of predictive research. However, these studies could not be definitively ruled out as having causal aims when assessed for inclusion. We additionally conducted a sensitivity analysis with stricter criteria for studies to be considered causal. This sensitivity analysis was not originally included in the original study pre-registration. More details of the sensitivity analysis can be found in the supplement **(Supplement: Sensitivity Analysis, Supplement: Protocol Deviations)**.

## Results

### Study Sample

After screening all 4,404 retrieved articles, our overall sample consisted of 250 articles (**Figure 2**). These articles included the following patient populations: stroke (n=192; 76.8%), multiple sclerosis (n=25; 10%), Parkinson’s disease (n=12; 4.8%), traumatic brain injury (n=4; 1.6%), Huntington’s disease (n=3; 1.2%), encephalitis (n=3; 1.2%), Guillain-Barré (n=3; 1.2%), and other neuropathologies (n=10; 4.0%) (**Supplement Table 3**). Two studies contained dual-pathology populations (hemorrhagic stroke and traumatic brain injury; ischemic stroke and acute myocardial infarction) and were each counted twice, once for each patient population.

**Figure 2.**
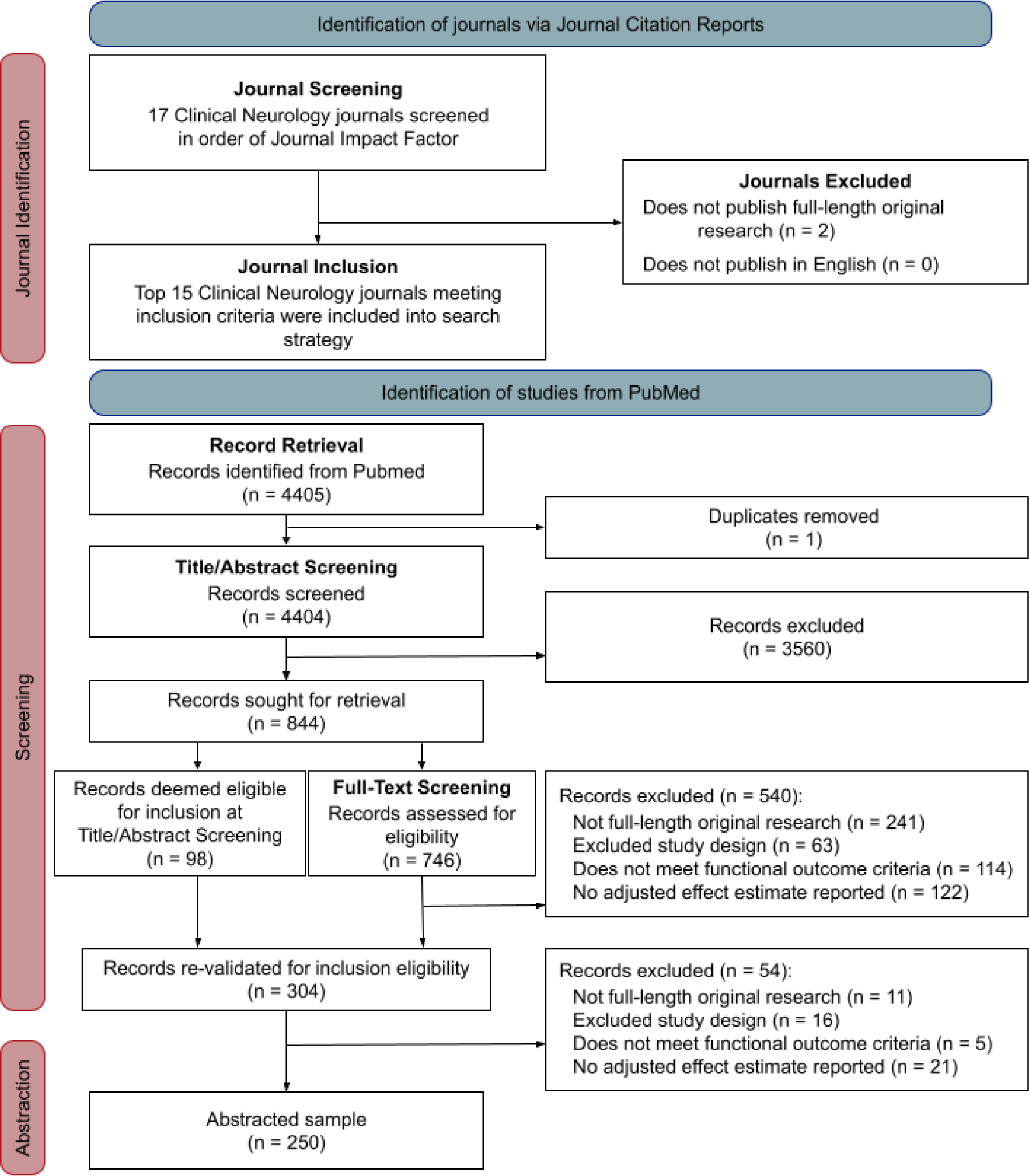
PRISMA diagram of the study selection. “Not full-length original research” indicates short-form research (e.g., brief report, research letter, without a Background, Methods, Results, or Discussion section, or not original research); “Excluded study design” indicates a prediction model, systematic review, or non-observational research; “Does not meet functional outcome criteria” indicates no outcome in the study meets our functional outcome criteria (see **Supplement: Definitions**); “No adjusted effect estimate reported” indicates no effect estimate for the functional outcome, or the effect estimate is not adjusted. Records that appeared to meet all inclusion criteria during title/abstract screening underwent another validation that all inclusion criteria were met.

### Prevalence of Grotta bars

In our overall sample (n = 250), 93 (37.2%) studies used at least one Grotta bars graph to report exposure-outcome relationships (**Figure 3**). Some included studies presented more than one Grotta bars graph, and the same functional outcome was visualized across all Grotta bars in these studies. The mRS was the most frequently visualized functional outcome (n=86; 92.4%), followed by the National Institutes of Health Stroke Scale (n=2; 2.2%), Guillain-Barré syndrome disability score (n=2; 2.2%), Pediatric Stroke Outcome Measure (n=1; 1.1%), Glasgow Outcome Scale Extended (n=1; 1.1%), and the Rankin Scale (RS) (n=1; 1.1%) **(Supplement: Table 4)**. Most studies using Grotta bars had a stroke patient study population (n=87; 93.5%), followed by encephalitis (n=3; 3.2%), Guillain-Barré (n=2; 2.2%), traumatic brain injury (n=1; 1.1%), and acute myocardial infarction (n=1; 1.1%).

**Figure 3.**
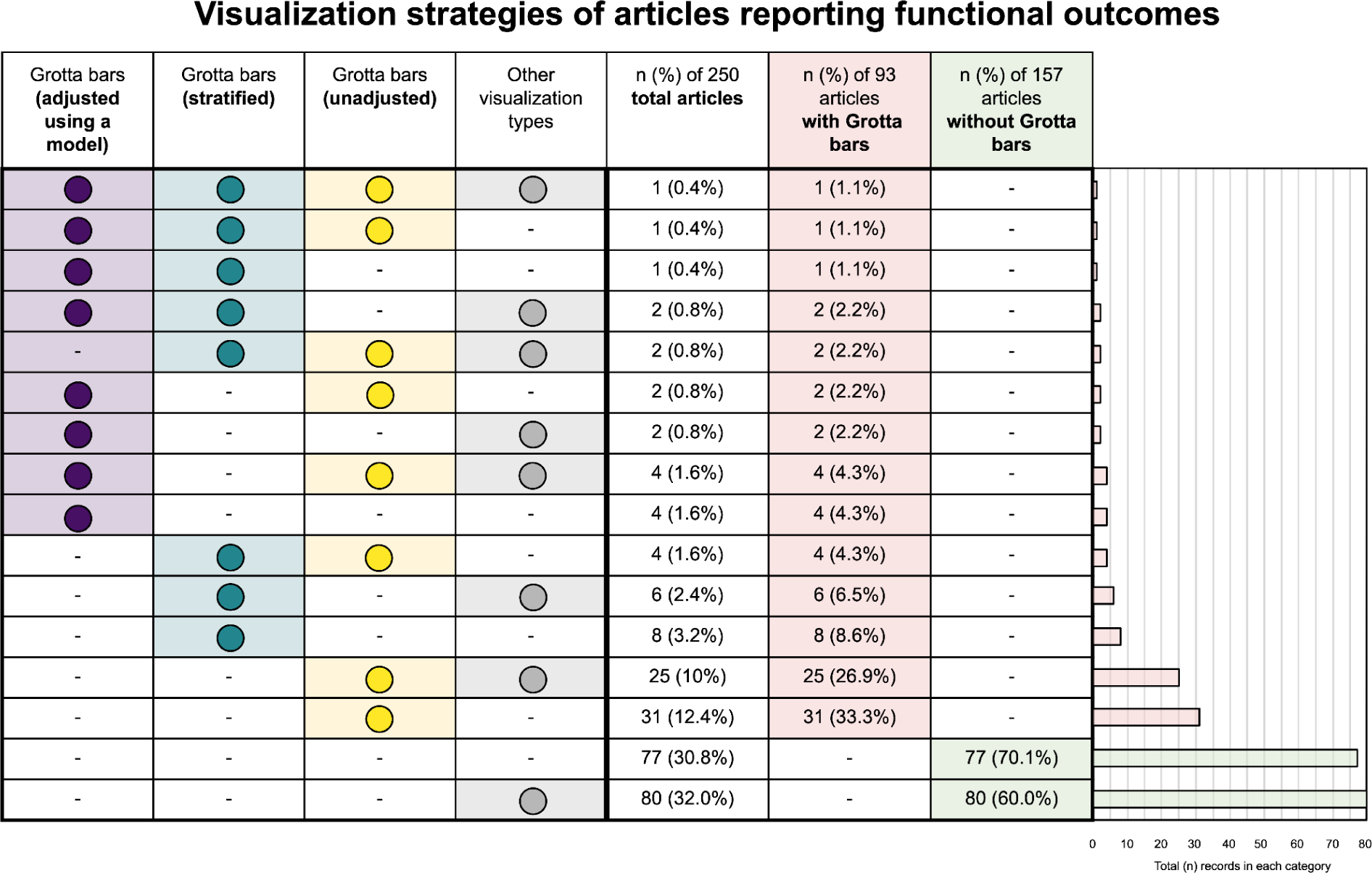
Functional outcome visualization in the overall sample. This table outlines whether each study contained Grotta bars that were unadjusted, stratified, and/or adjusted using a model to visualize functional outcomes, in addition to whether other visualization strategies were used to depict functional outcomes. The bar graph on the right of the figure depicts the number of studies using each combination of visualization strategies as an absolute count. An article was classified as having “other visualization types” if that article presented at least one visualization of the functional outcome that was not a Grotta bars graph.

### Adjusted Grotta bars

In accordance with our inclusion criteria, all studies in our sample reported effect estimates for one or more functional outcome(s) that were adjusted using a model-based approach. Among the 93 studies with Grotta bars, only 17 (18.3%) depicted Grotta bars that were adjusted using a model-based approach. The model-based adjustment strategies that were used in our sample included propensity score matching, (n=10; 58.8%), ordinal regression (n=6; 35.3%), and inverse probability weighting (n=1; 5.9%).

Furthermore, 25 of the 93 (26.9%) studies with Grotta bars visualized functional outcomes using stratified Grotta bars for at least one variable. Of these, 20 (80%) studies did not additionally present Grotta bars adjusted using a model. We present an overview of the adjustment strategies of the Grotta bars (unadjusted, stratified, and/or model-based adjustment), as well as whether articles used other visualization strategies, in **Figure 3**.

### Sensitivity Analysis

The results of the sensitivity analysis are very similar to the results of the main analysis **(Supplemental Materials: Sensitivity Analysis).**

## Discussion

We performed a meta-research study of observational, neurological research studies published in 15 top journals in 2020 and 2021 that present associations between an exposure and a functional outcome adjusted for confounding. Our study determined that stroke research, in particular, often used Grotta bars to visualize relationships between exposures and functional outcomes. We also identified isolated occurrences in encephalitis, Guillain-Barré, and traumatic brain injury research (as well as acute myocardial infarction in a population combined with a stroke). By design, every article included in our study appeared to control for confounding using a model-based method. However, more than 80% of studies reporting Grotta bars did not adjust Grotta bars using a model-based method. This indicates that generating adjusted Grotta bars reflective of their accompanying adjusted effect estimates is not common practice in observational neurological research.

### Utility of Grotta bars to depict functional outcomes

Grotta bars are useful in depicting ordinal functional outcomes by exposure groups at a high level of granularity. When the levels of the outcome scale are not collapsed (e.g., dichotomized or trichotomized), readers can readily identify the differences in the full outcome distributions. This feature is notable for patients and clinicians interested in observing the shift at a specific, clinically relevant level of granularity.^6^ We therefore recommend presenting all levels of the outcome scale on Grotta bars. This practice enhances the understandability of the graph for the reader, adheres to current guidelines for the statistical analysis of ordinal functional outcomes,^5^ and as a result, can help inform decision-making for clinicians and other stakeholders.^5,6^

Grotta bars can be misleading if they do not include the adjustments applied in the analysis.^4^ Unadjusted Grotta bars may be useful for descriptive purposes in observational studies to present the observed exposure-outcome associations. However, readers familiar with these bars from randomized trials may be tempted to improperly infer causality from the unadjusted Grotta bars in observational studies. For this reason, we advise authors of observational studies to present an adjusted version alongside unadjusted Grotta bars when aiming to answer causal questions. We believe this recommendation complements checklist item 16b of the Strengthening the reporting of observational studies in epidemiology (STROBE) guidelines, which states, “give unadjusted estimates and, if applicable, confounder adjusted estimates.”^19^

### Strategies to generate adjusted Grotta bars

Three reported methods of model-based confounding adjustment were used to create adjusted Grotta bars in our sample: ordinal outcome regression, propensity score matching, and inverse probability of treatment weighting. These adjustment strategies are summarized in **Figure 4**. Stratification by a variable other than the exposure was commonly encountered in Grotta bars both with and without model-based adjustment. Stratification alone, however, is likely not sufficient to address the threat of unmeasured confounding.

**Figure 4.**
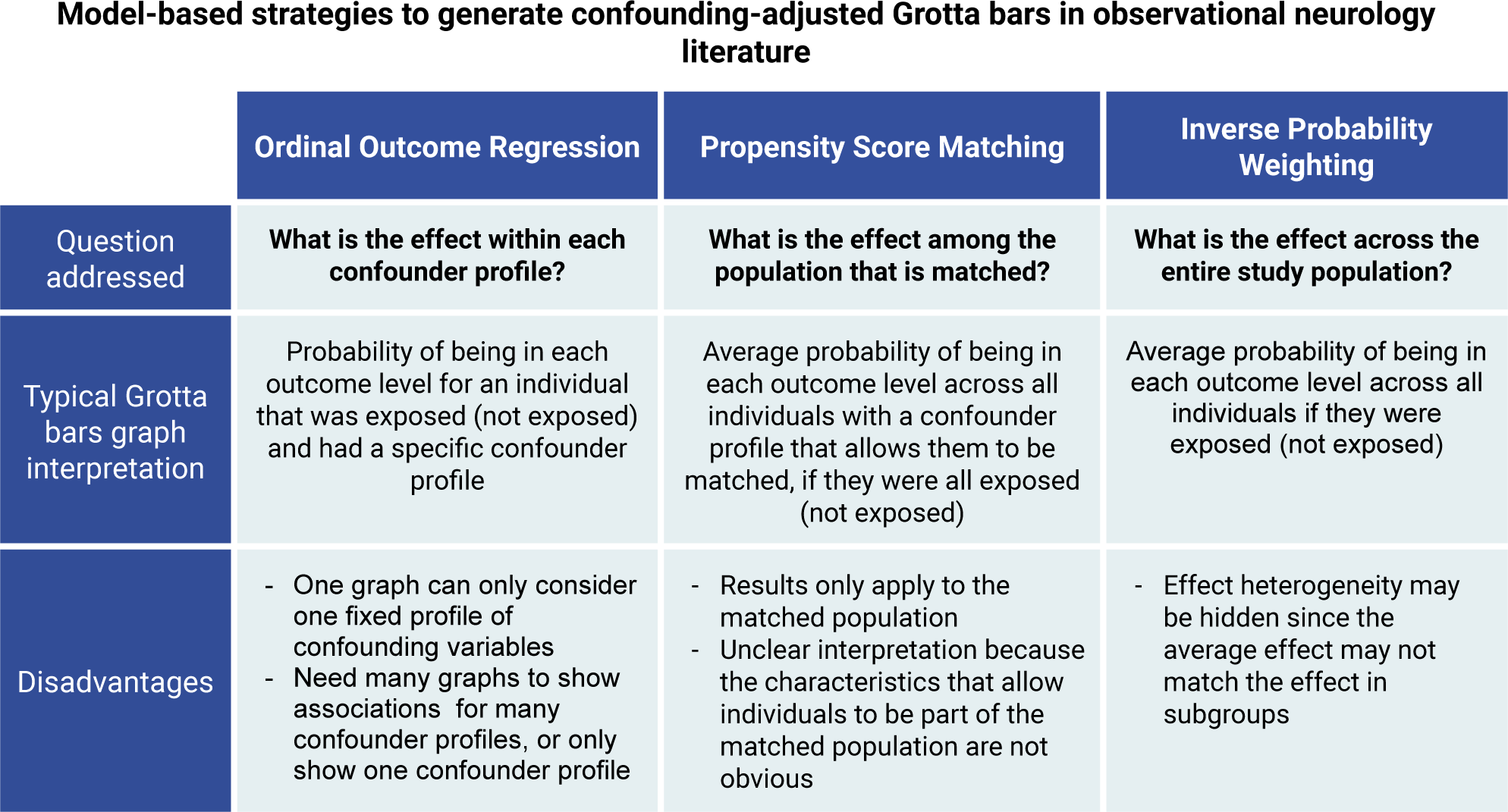
Summary of model-based adjustment methods identified in our sample of studies. Amongst our sample of 17 studies that depict Grotta bars adjusted using models, ordinal regression (n=6), propensity score matching (n=10), and inverse probability weighting (n=1) were the techniques used to adjust the visualizations.

Selecting an appropriate adjustment strategy is imperative to generate Grotta bars that are both interpretable and aligned with the target causal effect^20^ (**Figure 4**; **Figure 5**). In the same set of data with identical exposures, outcomes, and confounders, the application of different adjustment strategies can produce different results because the methods may target different causal effects.^20^ Researchers must start with a well-defined question; then select the appropriate adjustment strategy to estimate the causal effect targeted by their research question. This is essential to draw valid causal inferences from observational analyses.^4^

**Figure 5.**
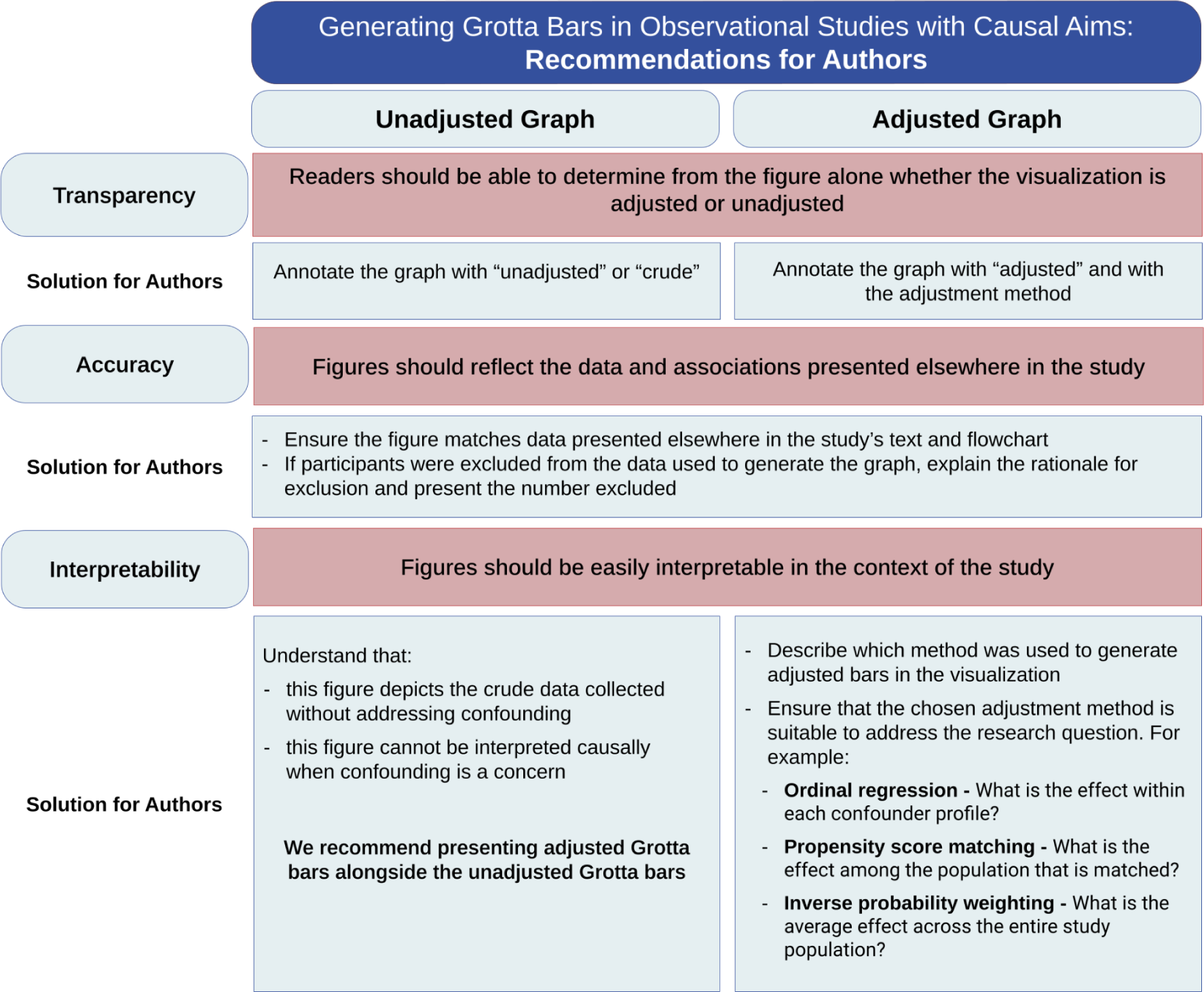
Recommendations for authors. We provide guidance for creating interpretable and transparent Grotta bars, based on the barriers limiting the interpretability of Grotta bars identified in our study.

Research designed to impact public health policies often aims to determine the causal effect across the entire study population.^10^ In this situation, inverse probability weighting is an adjustment strategy that is particularly suitable to remove confounding influence and generate corresponding adjusted Grotta bars.^4^ This strategy allows us to estimate the average (i.e., marginal) effect amongst all individuals within a given study population, which is typically the effect reported in randomized controlled trials.^10^ Rohmann *et al.* provide a detailed tutorial on how to build and interpret Grotta bars by applying inverse probability weighting, using stroke registry data.^4^ Our results suggest that this technique is underutilized in observational studies published in top clinical neurology journals.

### Causal conclusions from observational data

While unmeasured confounding can be a major obstacle for causal inference in observational settings, observational datasets are useful when the experimental studies needed to answer a research question are unethical or unfeasible.^21^ Effect estimates and visualizations depicting results from observational studies have the potential to be endowed with a causal interpretation if careful considerations are made in the design and analysis of an observational study.^10^ While it is an important first step, the presentation of adjusted Grotta bars from observational data does not guarantee a straightforward causal interpretation (**Figure 6**).^10,22^

**Figure 6.**
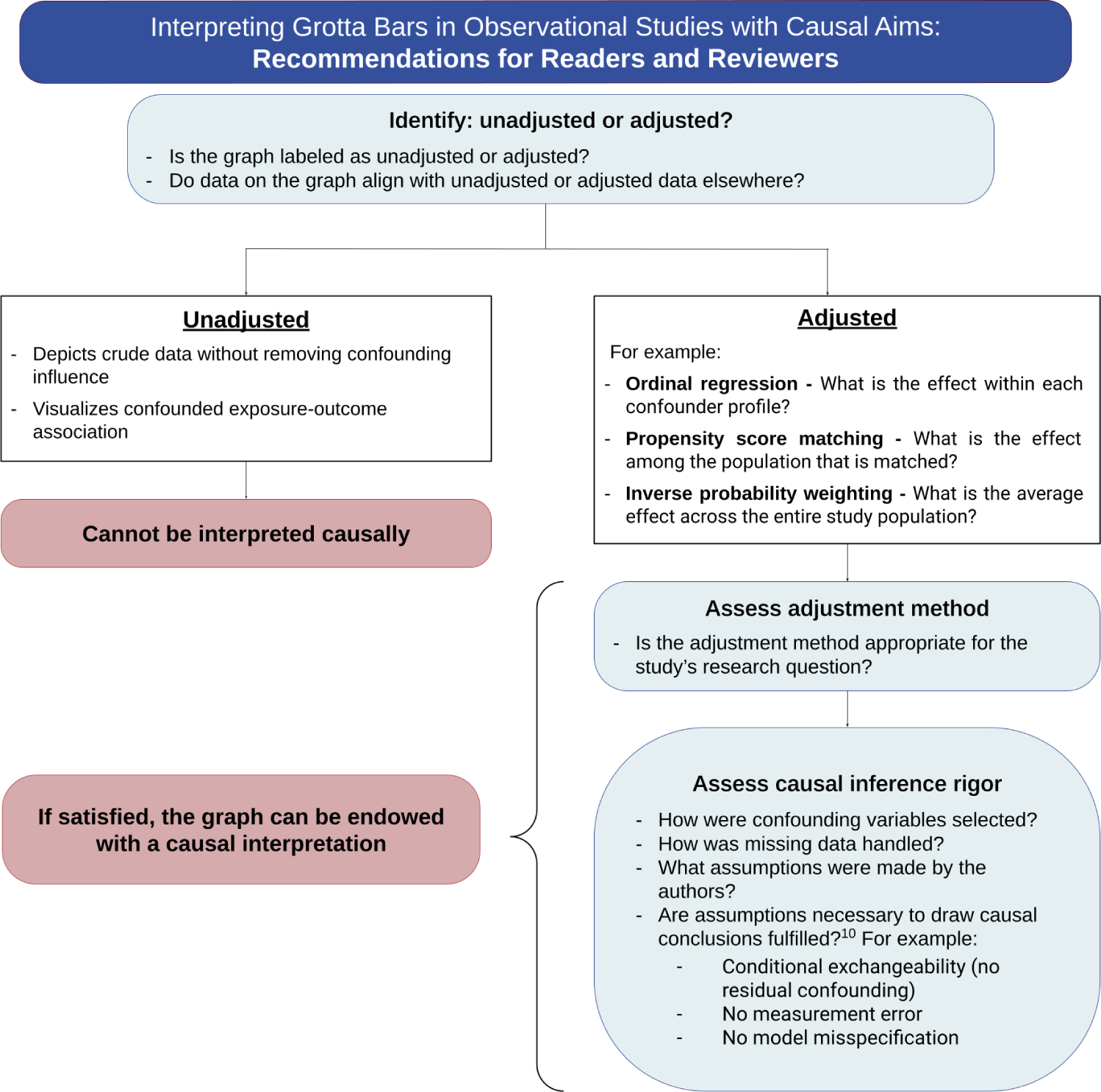
Recommendations for readers and reviewers. Guidance for interpreting Grotta bars when the association between exposure and functional outcome is adjusted in the analysis. We recommend that readers have a nuanced approach when interpreting Grotta bars in observational neurological research depending both on whether adjustment is present and which adjustment method is used.

### Limitations

This study has several limitations. First, our sample only included articles from the top-ranked subset of journals from the Journal Citation Reports “Clinical Neurology” category. Additionally, only articles written in English were included in our sample. Our results may not be generalizable to articles published in other journals. Second, only the main texts of articles were examined. It is possible that supplementary materials for publications containing Grotta bars went unaccounted for in our study, and that these Grotta bars were generated differently than those present in the main article. Third, we did not attempt to assess the appropriateness or rigor of the methods applied to individual studies included in our sample. As such, we cannot conclude whether any of the adjusted Grotta bars in our sample can be interpreted causally. Finally, we aimed to only include publications with causal aims in our study. Our assessment of whether individual studies had causal aims may not always reflect the authors’ intentions. Causal aims are subjective and are known to be inconsistently reported.^15,16^ It is therefore possible that we excluded studies with causal aims and conversely, that studies without causal aims were included in our sample.

We emphasize that this article specifically is focused on confounding. Unadjusted Grotta bars may also represent biased exposure-outcome association if other types of biases (e.g., selection bias) are present. While confounding is a major concern in observational studies, we encourage readers to carefully consider other sources of biases when they perform causal inference analyses.^10,23,24^

## Conclusion

In conclusion, our study shows that mismatches between the presentation of adjusted effect estimates and their accompanying Grotta bars are common in observational neurology research focusing on functional outcomes. These figures can be misleading as unadjusted Grotta bars created from observational data represent confounded exposure-outcome associations and cannot be interpreted causally. If graphs depict observed functional outcome distributions by exposure groups, readers must recognize that any observable distributional difference may be explained by confounding and may not match the adjusted effect estimates. If a Grotta bars graph is adjusted, a causal interpretation may be possible depending on the research aims and whether rigorous causal inference methods were applied in the study.

Visualizations have great potential to enhance the reader’s understanding of study results.^25^ Authors should generate visualizations that reflect a study’s result to clearly and informatively present research findings.^26^ Grotta bars are a very effective tool for visualizing the strength and direction of ordinal distributional shifts, and are very popular in randomized stroke trials. We believe this graphical tool can also be useful in observational studies examining effects on functional outcomes. We recommend that both an unadjusted Grotta bars graph and an adjusted version be presented together when adjusted effects for functional outcomes are presented in a study.

## Supporting information

Supplement: Definitions

Supplement: Protocol Deviations

Supplement: Sensitivity Analysis

## Data Availability

All protocols were pre-registered on our Open Science Framework (RRID:SCR_003238) repository which also contains full protocols, protocol modifications, and a full abstraction table. Data and code are available on our GitHub repository.

https://github.com/meghanrforrest/

https://osf.io/w78mh/

## Declarations

### Ethical approval

Ethical approval was not sought for the present study because this was a study of published literature.

### Informed consent

Informed consent was not sought for the present study because there were no participants in this study of published literature.

### Guarantor

MRF and JLR

### Contributorship

MRF: Conceptualization, data curation, formal analysis, investigation, methodology, project administration, software, validation, visualization, writing - original draft preparation, writing - review and editing; TLW: methodology, visualization, writing - reviewing and editing; ESL: investigation, writing - review and editing; ETC: investigation, writing - review and editing; EF: investigation, writing - review and editing; LJ: methodology, writing - review and editing; MP: conceptualization, methodology, supervision, visualization, writing - review and editing; JLR: conceptualization, formal analysis, funding acquisition, methodology, project administration, resources, supervision, visualization, writing - original draft preparation, writing - review and editing.

## Acknowledgements

We would like to thank Camila Victoria-Quilla Baselly Heinrich for her advice regarding the data management practices. We thank Nadja Wülk for her proofreading and support of this project as MRF’s master’s thesis, as well as Fiona Morrison for additional grammatical proofreading support.

