## Supplement: Table 3 for "Use of stacked proportional bar graphs (“Grotta bars”) to visualize functional outcome distributions in observational neurology research"

**Table 3. Neurological conditions in the sample**

| | Total (N=252) | No Grotta bars (N=158) | Unadjusted Grotta bars or Stratified Grotta bars only (N=77) | At least one model-based adjusted Grotta bars graph (N=17) |
|---|---|---|---|---|
| Acute myocardial infarction * | 1 (0%) | 0 (0%) | 1 (1%) | 0 (0%) |
| Dementia | 1 (0%) | 1 (1%) | 0 (0%) | 0 (0%) |
| Encephalitis | 3 (1%) | 0 (0%) | 3 (4%) | 0 (0%) |
| Guillain-Barré | 3 (1%) | 1 (1%) | 2 (3%) | 0 (0%) |
| Huntington’s disease | 3 (1%) | 3 (2%) | 0 (0%) | 0 (0%) |
| Inclusion body myositis | 1 (0%) | 1 (1%) | 0 (0%) | 0 (0%) |
| Meningitis | 2 (1%) | 2 (1%) | 0 (0%) | 0 (0%) |
| Multiple sclerosis | 25 (10%) | 25 (16%) | 0 (0%) | 0 (0%) |
| Neonatal hypoxic ischemic encephalopathy | 1 (0%) | 1 (1%) | 0 (0%) | 0 (0%) |
| Parkinson's disease | 12 (5%) | 12 (8%) | 0 (0%) | 0 (0%) |
| Seizures | 2 (1%) | 2 (1%) | 0 (0%) | 0 (0%) |
| Spinal cord injury | 1 (0%) | 1 (1%) | 0 (0%) | 0 (0%) |
| Spinocerebellar ataxia type 2 | 1 (0%) | 1 (1%) | 0 (0%) | 0 (0%) |
| Stroke (hemorrhagic and ischemic) | 9 (4%) | 7 (4%) | 2 (3%) | 0 (0%) |
| Stroke (hemorrhagic) † | 24 (10%) | 18 (11%) | 4 (5%) | 2 (12%) |
| Stroke (ischemic) * | 159 (63%) | 80 (51%) | 64 (83%) | 15 (88%) |
| Traumatic brain injury † | 4 (2%) | 3 (2%) | 1 (1%) | 0 (0%) |

* One occurrence of acute myocardial infarction and ischemic stroke in a combined population

† One occurrence of stroke (hemorrhagic) and traumatic brain injury in a combined population
