## Supplement: Table 4 for "Use of stacked proportional bar graphs (“Grotta bars”) to visualize functional outcome distributions in observational neurology research"

**Table 4. Functional outcomes included in the sample.**

| | Total (N=359) |
|---|---|
| **Functional outcome** | |
| Barthel Index | 5 (1%) |
| Bayley III | 1 (0%) |
| Expanded Disability Status Scale | 25 (7%) |
| Glasgow Outcome Scale | 4 (1%) |
| Glasgow Outcome Scale - Extended | 3 (1%) |
| Glasgow Outcome Scale - Extended (pediatric) | 1 (0%) |
| Gross Motor Function Classification System | 1 (0%) |
| Guillain-Barré Syndrome Disability Scale | 3 (1%) |
| Inclusion Body Myositis-Functional Rating Scale | 1 (0%) |
| Inflammatory Rasch-Built Overall Disability Scale | 1 (0%) |
| International Cooperative Ataxia Rating Scale | 1 (0%) |
| International Standards for Neurological Classification of Spinal Cord Injury | 1 (0%) |
| Inventory of Non-Ataxia Signs | 1 (0%) |
| Katz Index | 1 (0%) |
| King's Outcome Scale for Childhood Head Injury | 1 (0%) |
| Lawton–Brody scale | 1 (0%) |
| Medical Research Council Sum Score | 1 (0%) |
| Modified National Institutes of Health Stroke Scale | 1 (0%) |
| Modified Rankin Scale | 179 (50%) |
| Multiple Sclerosis Severity Score | 2 (1%) |
| Multiple Sclerosis Severity Score (pediatric) | 1 (0%) |
| National Institutes of Health Stroke Scale | 90 (25%) |
| National Institutes of Health Stroke Scale (pediatric) | 2 (1%) |

| | |
|---|---|
| Pediatric Cerebral Performance Category Scale | 1 (0%) |
| Pediatric Stroke Outcome Measure | 3 (1%) |
| Rankin Scale | 1 (0%) |
| Rasch-Built Overall Built Disability Score | 1 (0%) |
| Scale for Assessment and Rating of Ataxia | 1 (0%) |
| Scales for Outcomes in Parkinson's Disease | 1 (0%) |
| Spinal Cord Independence Measure, Version III | 1 (0%) |
| Stroke Impact Scale 16 | 2 (1%) |
| Stroke Levity Scale | 1 (0%) |
| The Unified Huntington's Disease Rating Scale - Total Functional Capacity | 1 (0%) |
| The Unified Huntington's Disease Rating Scale - Total Motor Score | 1 (0%) |
| The Warner Initial Developmental Evaluation of Adaptive and Functional Skills | 1 (0%) |
| Unified Huntington's Disease Rating Scale | 2 (1%) |
| Unified Parkinson's Disease Rating Scale (composite) | 2 (1%) |
| Unified Parkinson's Disease Rating Scale-III | 11 (3%) |
| Utility-Weighted Modified Rankin Scale | 2 (1%) |
