## Supplement: Definitions for "Use of stacked proportional bar graphs (“Grotta bars”) to visualize functional outcome distributions in observational neurology research"

**Observational study** – Non-randomized studies with a control group

- Inclusion examples
    - Cohort study
    - Case-control study
    - Cross-sectional study
    - Case-cohort study
    - Nested case control study
- Exclusion examples
    - randomized controlled trial
    - Cluster randomized controlled trial
    - Other clinical trial
    - Case study
    - Case series
    - Meta-analysis
    - Self-controlled designs
        - Case crossover
        - Self-controlled case series
        - Sequence symmetry analysis
    - Prediction model studies
        - Internal validation and/or external validation specified and/or
        - Authors report that they built a prediction model

    <u>Note on randomized controlled trials:</u>

    - We acknowledge that secondary analyses of a randomized dataset may fall in the category of an “observational study” if the researchers do not use the randomization variable in the randomized controlled trial as the exposure in the secondary analysis/post-hoc analysis. We are making the following decision on how to handle this situation because we believe it will have a high sensitivity of capturing randomized controlled trial data used observationally:
        1. If the Abstract notes that a registry following or from a randomized controlled trial is the source of the data, then the study will be considered an *observational study*
        2. If the Abstract identifies that clinical trial/randomized controlled trial data were used without an indication for adjustment for the originally randomized exposure variable in the Abstract (which would be needed in an observational analysis of data originally collected in a trial), then the study <u>*will not*</u> be *considered an observational study*. We assume that authors will define in the Abstract how they handled the randomization in order for the data to be analyzed in an observational manner

**Functional outcomes** (used as either primary or secondary endpoints, not as a covariate in analysis) must fulfill *each* of the following criteria:

- ☐ Ordinal scale
- ☐ Measures global physical functional ability (i.e. not only hand or arm or leg dexterity or strength, not only a measure of cognition such as GCS)
- ☐ The outcome scale is evaluated by an external assessor and is not a self-evaluation (e.g. survey)
- ☐ The outcome scale is not used for disease screening

*Please refer to Pre-Registration pg. 16 for examples of functional outcome inclusion decisions*

**Grotta bars** comparing exposure groups must meet *each* of the following criteria:

- ☐ X-axis has a range from 0% - 100%, or is a proportion from 0 to 1
- ☐ Components (segments) of each individual bar sum to 100%
- ☐ Bars are broken down into smaller segments based on an ordinal scale
- ☐ Bars can be vertical or horizontal
- ☐ Bars have more than 2 segments
- ☐ Lines connecting bars may or may not be present

- For inclusion and exclusion examples with justification, please refer to *Pre-Registration pg. 17-25*

**Effect measures** “indexes that summarize the strength of the link between exposures and outcomes” [27]

- Examples:
    - Odds ratio
    - Risk ratio
    - Risk difference
    - Rate Difference
    - Hazard ratio
    - Incidence rate ratio
    - Number needed to treat
