## Supplement: Protocol Deviations for "Use of stacked proportional bar graphs (“Grotta bars”) to visualize functional outcome distributions in observational neurology research"

### Major Deviations

Abstraction

- The first three batches of randomized papers (n=5) were distributed to all three second reviewers (EL, ET, EF) after they underwent abstraction by MRF, who managed all data. Once all four individuals had independently completed each abstraction batch, all four abstractors met to review decision discrepancies and altogether, develop a "gold standard" for how the questions should be answered according to the protocol. The fourth batch increased in sample size (n=10). All four reviewers met once again for a "gold standard" development meeting to review discrepancies, after which all abstractors reported feeling increased confidence with their understanding of the protocol. A Fischer's Kappa was calculated, which resulted in 86.5% inter-rater agreement despite some contingency questions having discrepancies. This number was deemed sufficient for the second abstractor to receive their own sample starting with abstraction batch 5.

Abstraction form and variables

- We added the following variables to our abstraction (for the purpose of refining our sample for likelihood of causal intentions by the authors):
    - An receiver operator characteristic curve and/or area under the curve interpreted pertaining to the functional outcome
    - The types of adjusted effect estimates present in the study, in relation to the functional outcome
    - Whether the authors explicitly describe their intention for including covariates into their model is to control/adjust for confounding, or similar
    - We added the following variable to the abstraction
        - Functional outcome is visually depicted in a different type of figure that is not a stacked proportional bar graph (yes/no)
- We did not allow stratified graphs without further adjustment to qualify as "adjusted", because this is only conditioning on the stratified variable. We instead differentiated between "model adjusted" and "stratified" Grotta bars
- We did not generate "a bar graph visualization of functional outcomes which are presented in Grotta bars" because an overwhelming majority were only depicting mRS
- We refined our sample beyond the original inclusion criteria because the decision of whether studies have causal intentions was often not straightforward.
- No confidence intervals were calculated, as our sample includes each study meeting all inclusion requirements

### Minor Deviations

Screening

- A fourth screener (EF) was added for full-text screening.

Abstraction

- A fourth abstractor (EF) was added to the abstraction team. MRF screened 100% of texts. A random sample of papers was distributed in batches to each other abstractor (EL, ET, EF) for a second independent abstraction according to the abstractor's availability
- In addition to looking at the Grotta bars Figure or the Grotta bar Figure's description for information about adjustment, abstractors additionally searched for the Figure name in the paper (CTRL + F) to see if there was any additional information about adjustment relating to the Figure

Abstraction form and variables

- We did not use Google Forms for our abstraction. Instead, we used MS Forms because

1) the MS Forms branching feature was easier to devise when constructing the survey
2) MS Forms is directly compatible with MS Excel

- The introduction page did not only ask whether the article is full-length original research, but asked abstractors to confirm that the article meets all inclusion criteria. Abstraction ended if the article was deemed to not meet all inclusion criteria
- The following study designs were added to the list of pre-specified study designs:
    - Nested cohort
- The following functional outcomes were added to the list of pre-specified functional outcomes:
    - Rankin Scale (RS)
    - EDSS
    - UPDRS-III
    - UPDRS (composite)
    - Balance Evaluation Systems Test (BESTest)
- The following neurological condition was added to the list of pre-specified neurological conditions:
    - Multiple sclerosis

Open Science Sharing

- Data and code available on our GitHub repository rather than OSF to facilitate analysis and code editing

**Protocol Clarifications**

Inclusion/Exclusion criteria

- In the Pre-Registration, we state in our last inclusion criteria: “Contains a confounding-adjusted effect estimate (Abstract, main-text Results section, or main-text table).” Reviewers clarified that the effect estimate of the *functional outcome* must be adjusted to be eligible for inclusion

Abstraction

- If the Grotta bar was not explicitly identified as adjusted, but was presented either with an adjusted effect estimate directly in the Figure, in the Figure’s description, or in the text in reference to the Figure, abstractors were instructed to look for functional outcome data to make a well-informed decision regarding whether the figure is adjusted or unadjusted. Presenting an adjusted effect estimate in the Figure, in the Figure’s description, or in the text in reference to the Figure is not sufficient to decide that the Figure is adjusted.
