## Supplement: Sensitivity Analysis for "Use of stacked proportional bar graphs (“Grotta bars”) to visualize functional outcome distributions in observational neurology research"

**Methods**

To be included in the sensitivity analysis sample with stricter criteria for evidence of causal aims, studies must meet all of the following criteria: (1) reported one or more coefficients for the functional outcome that were transformed into an effect measure (such as a risk ratio or odds ratio)[28]; (2) did not feature a receiver operator characteristic curve and/or an area under the curve metric in the interpretation of an exposure-outcome relationship; and (3) specified that inclusion of covariates into the statistical model is specifically intended to address confounding.[17,18,29,30]

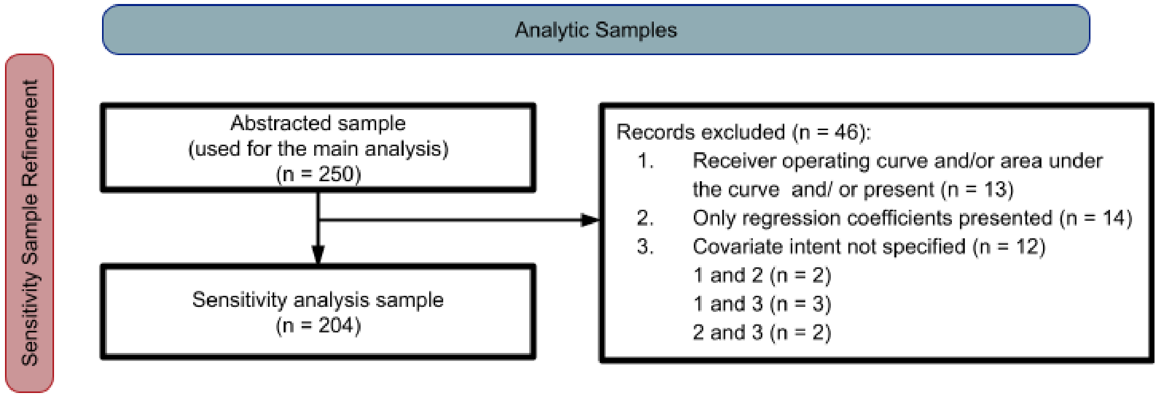


**Results**

In total, 204 studies were included in the sensitivity analysis sample. Of these, 88 (43.1%) contained at least one Grotta bars graph, most of which lacked a version of the graph that was adjusted with a model-based adjustment strategy (n = 71; 80.7%).
