## Supplement: Table 1 for "Use of stacked proportional bar graphs (“Grotta bars”) to visualize functional outcome distributions in observational neurology research"

### Supplementary Files

**Table 1.** Results from journal screening.

| Rank | Name | ISSN | eISSN | Exclusion Reason |
|---|---|---|---|---|
| 1 | Lancet Neurology | 1474-4422 | 1474-4465 | - |
| - | Nature Reviews Neurology | 1759-4758 | 1759-4766 | does not publish original research |
| 2 | JAMA Neurology | 2168-6149 | 2168-6157 | - |
| 3 | Alzheimer’s & Dementia | 1552-5260 | 1552-5279 | - |
| 4 | Acta Neuropathologica | 0001-6322 | 1432-0533 | - |
| 5 | Brain | 0006-8950 | 1460-2156 | - |
| 6 | Journal of Neurology Neurosurgery and Psychiatry | 0022-3050 | 1468-330X | - |
| 7 | Neuro-Oncology | 1522-8517 | 1523-5866 | - |
| 8 | Neurology | 0028-3878 | 1526-632X | - |
| 9 | Psychiatry and Clinical Neurosciences | 1323-1316 | 1440-1819 | - |
| - | SLEEP MEDICINE REVIEWS | 1087-0792 | 1532-2955 | does not publish original research |
| 10 | Neurology-Neuroimmunology & Neuroinflammatory | 2332-7812 | 2332-7812 | - |
| 11 | Annals of Neurology | 0364-5134 | 1531-8249 | - |
| 12 | Stroke | 0039-2499 | 1524-4628 | - |
| 13 | Stroke and Vascular Neurology | 2059-8688 | 2059-8696 | - |
| 14 | Movement Disorders | 0885-3185 | 1531-8257 | - |
| 15 | Brain Stimulation | 1935-861X | 1876-4754 | - |

These results were inserted into items 1 (ISSN) and 2 (eISSN) of the search strategy (Supplementary Table 4). No discrepancies arose during journal screening.
