## Supplement: Table 2 for "Use of stacked proportional bar graphs (“Grotta bars”) to visualize functional outcome distributions in observational neurology research"

**Table 2.** Complete search strategy

| Search Strategy, Pubmed: November 08, 2022 | | |
|---|---|---|
| 1 | 1474-4422[ta] OR 2168-6149[ta] OR 1552-5260[ta] OR 0001-6322[ta] OR 0006-8950[ta] OR 0022-3050[ta] OR 1522-8517[ta] OR 0028-3878[ta] OR 1323-1316[ta] OR 2332-7812[ta] OR 0364-5134[ta] OR 0039-2499[ta] OR 2059-8688[ta] OR 0885-3185[ta] OR 1935-861X[ta] | ISSN |
| 2 | 1474-4465[ta] OR 2168-6157[ta] OR 1552-5279[ta] OR 1432-0533[ta] OR 1460-2156[ta] OR 1468-330X[ta] OR 1523-5866[ta] OR 1526-632X[ta] OR 1440-1819[ta] OR 2332-7812[ta] OR 1531-8249[ta] OR 1524-4628[ta] OR 2059-8696[ta] OR 1531-8257[ta] OR 1876-4754[ta] | eISSN |
| 3 | #1 OR #2 | |
| 4[13] | ((cohort[all] OR (control[all] AND study[all]) OR (control[tw] AND group*[tw]) OR epidemiologic studies[mh] OR program[tw] OR clinical trial[pt] OR comparative stud*[all] OR evaluation studies[all] OR statistics as topic[mh] OR survey*[tw] OR follow-up*[all] OR time factors[all] OR ci[tw]) NOT ((animals[mh:noexp] NOT humans[mh:noexp]) OR comment[pt] OR editorial[pt] OR review[pt] OR meta analysis[pt] OR case report[tw] OR consensus[mh] OR guideline[pt] OR history[sh])) | Search filter to identify controlled non-randomized studies<br><br>Sensitivity: 92.42%<br>Specificity: 79.67%[13] |
| 5 | 2020[PDAT]:2021[PDAT] | |
| 6 | #3 AND #4 AND #5 | |
